# Mindfulness mediates depressive symptom improvement during heated yoga: a secondary analysis of a randomized controlled trial

**DOI:** 10.64898/2026.07.01.26353530

**Authors:** Daniel I. Copeland, Naoise Mac Giollabhui, Louisa Sylvia, Doga Cetinkaya, Serene J. Puzak, Lindsey B. Hopkins, Chris C. Streeter, Bettina B. Hoeppner, Lisa Uebelacker, Jill Koontz, Simmie Foster, Christina Dording, Albert Yeung, Lauren B. Fisher, Cristina Cusin, Felipe A. Jain, Paola Pedrelli, Grace A. Ding, Heather Raslan, Ashley E. Mason, Paolo Cassano, Darshan H. Mehta, Charles L. Raison, Christina Sauder, Karen K. Miller, Brian W. Anthony, Maurizio Fava, David Mischoulon, Maren B. Nyer

**Affiliations:** Department of Psychiatry, Massachusetts General Hospital, Boston, Massachusetts, United States of America; Department of Psychiatry, Harvard Medical School, Boston, Massachusetts, United States of America; Private Practice, San Francisco, California, United States of America; Department of Psychiatry and Neurology, Boston University School of Medicine, Boston, Massachusetts, United States of America; Alpert Medical School, Brown University, Providence, Rhode Island, United States of America; Butler Hospital, Providence, Rhode Island, United States of America; Blueprint Wellness, LLC, Wakefield, Massachusetts, United States of America; Osher Center for Integrative Health, University of California, San Francisco (UCSF), San Francisco, California, United States of America; Department of Psychiatry, University of California, San Francisco (UCSF), San Francisco, California, United States of America; Benson-Henry Institute for Mind Body Medicine, Massachusetts General Hospital, Boston, Massachusetts, United States of America; Osher Center for Integrative Medicine, Brigham and Women’s Hospital, Boston, Massachusetts, United States of America; Neuroendocrine Unit, Department of Medicine, Harvard Medical School, Boston, Massachusetts, United States of America; Department of Psychiatry, University of Wisconsin–Madison, Madison, Wisconsin, United States of America; Institute for Medical Engineering and Science (IMES), Massachusetts Institute of Technology, Cambridge, Massachusetts, United States of America; Department of Human Development and Family Studies, School of Human Ecology, University of Wisconsin–Madison, Madison, Wisconsin, United States of America

## Abstract

**Background:** Despite growing evidence that lifestyle interventions reduce depressive symptoms, the psychological mechanisms underlying these effects remain poorly understood. This study examined whether mindfulness and rumination mediate the antidepressant effects of heated yoga (HY), a multicomponent intervention combining physical activity, attentional training, and thermoregulatory stress.

**Methods:** This prespecified secondary mediation analysis builds on a randomized controlled trial in which 80 adults with moderate-to-severe depression (IDS-CR >23) were randomized to 8 weeks of twice-weekly HY or waitlist control [1]. Subsamples with complete mediator data contributed to rumination (*n* = 56) and mindfulness (*n* = 60) models. Causal mediation analyses with 10,000 bootstrap resamples estimated indirect effects on Week 8 depression severity via Week 4 mediator changes. Sensitivity analyses assessed unmeasured confounding required to nullify observed effects. ClinicalTrials.gov: NCT02607514.

**Results:** As previously reported, HY produced significantly greater IDS-CR reductions at Week 8 versus controls (*p* < .001). HY was associated with decreased rumination (*p* < .01) and increased mindfulness (*p* < .001) at Week 4. Increased mindfulness was statistically consistent with mediating depressive symptom reductions (ACME: −2.71, 95% CI [−5.42, − 0.99]), whereas decreased rumination was not (ACME: −2.41, 95% CI [−6.28, 0.43]). Results were resilient to sensitivity analyses.

**Conclusion:** In this RCT of a behavioral lifestyle intervention, mindfulness but not rumination emerged as a statistically significant mediator of depressive symptom reductions, identifying mindfulness as a key candidate mechanism through which multicomponent lifestyle interventions may exert antidepressant effects and suggesting a target for optimizing behavioral treatments for depression.

## Introduction

Depression is a leading cause of disability worldwide, affecting approximately 280 million individuals [2]. Although psychotherapy and antidepressant medications remain the most widely prescribed treatments, response rates are modest (up to 48% and 54%, respectively) [3, 4], and medication side effects including cognitive impairment, weight gain, and fatigue limit long-term adherence [5]. These treatment gaps have motivated growing interest in behavioral lifestyle interventions as adjunctive or standalone strategies for depression.

### Behavioral lifestyle interventions for depression

Lifestyle interventions, including aerobic exercise, dietary modification, sleep optimization, and mind-body practices, are increasingly recognized as first-line adjuncts in national depression guidelines [6, 7]. The behavioral medicine framework conceptualizes these interventions as targeting depression through multiple pathways: behavioral activation (increasing engagement with rewarding activities), physiological regulation (normalizing stress-responsive systems), and cognitive-affective change (altering maladaptive patterns of attention and thought) [8, 9]. However, the specific psychological and behavioral mechanisms through which lifestyle interventions reduce depressive symptoms remain insufficiently characterized, limiting efforts to optimize and personalize these treatments.

Yoga exemplifies a multicomponent behavioral intervention that simultaneously engages physical, cognitive, and regulatory processes. Studies suggest that yoga reduces rumination, anxiety, and depressive symptoms while improving psychological well-being in both clinical and sub-clinical populations [10–14]. The non-judgmental attention, self-acceptance, and breath regulation cultivated in yoga practice are hypothesized to reduce depression severity through behavioral mechanisms including enhanced present-moment awareness, reduced experiential avoidance, and improved distress tolerance [15–17].

### Heated yoga as a multicomponent behavioral intervention

Bikram yoga, also referred to as “heated yoga” (HY), combines traditional yoga practices with the added physiological stressor of ambient heat. HY is typically performed in an environment heated to 105°F (40.5°C), creating a multicomponent intervention that integrates: (1) physical activity through a standardized posture sequence; (2) attentional training requiring sustained focus on proprioceptive and interoceptive signals; (3) thermoregulatory stress from environmental heat exposure; and (4) behavioral regulation through managing discomfort and maintaining engagement during challenging conditions. Exposure to heated environments can induce whole-body hyperthermia (WBH), which has independent evidence supporting antidepressant effects through normalization of thermoregulatory cycling and modulation of serotonergic and interleukin-6 signaling pathways [18–22].

In the parent randomized controlled trial (RCT) for the present analysis, HY significantly reduced depressive symptoms compared to waitlist controls [1]. This is a prespecified secondary analysis of that trial, designed to identify the psychological mechanisms through which HY exerts its antidepressant effects.

### Candidate mediators: mindfulness and rumination

Mindfulness-based interventions (MBIs) have demonstrated efficacy in reducing depressive symptoms across multiple meta-analyses [23–26]. Theoretical models propose that mindfulness reduces depression through decentering (metacognitive distancing from negative thoughts), reduced experiential avoidance, improved emotion regulation, and enhanced self-compassion [27, 28]. Mindfulness was selected as a candidate mediator because HY requires sustained present-moment attention to physical sensation and discomfort in a heated environment, directly paralleling the attentional training cultivated in formal mindfulness meditation. The Five Facet Mindfulness Questionnaire (FFMQ), although originally developed in MBSR research, has been used in prior yoga studies and captures trait mindfulness dimensions relevant to yoga practice [29–31].

Rumination was selected as a candidate mediator based on stress-reactive rumination models [32, 33] and the cognitive model of depression, which posits that perseverative negative self-referential thinking maintains depressive episodes.

Interventions that disrupt rumination may exert antidepressant effects through an independent behavioral mechanism rather than merely reflecting symptom improvement. HY may reduce rumination by redirecting attention from self-referential thought to immediate physical experience, functioning as a form of behavioral redirection.

### Study aims

The aim of this paper is to examine two hypotheses:

1. Decrease in rumination scores at Week 4 will mediate the depressive symptom reduction of HY observed at Week 8.
2. Increase in mindfulness scores at Week 4 will mediate the depressive symptom reduction of HY observed at Week 8.

## Materials and methods

This study (ClinicalTrials.gov: NCT02607514) was approved by the Mass General Brigham Institutional Review Board (IRB Protocol Number 2016P000704). All subjects signed an IRB-approved informed consent statement. We followed the CLARIFY guidelines for reporting yoga research [34] and the CONSORT guidelines for reporting randomized trials [35] in describing our study design, results, and other relevant information. Participant flow through the study is detailed in the primary trial report [1].

### Procedure and randomization

A total of 80 participants were randomized to either 8 weeks of at least twice-weekly, 90-minute heated yoga sessions at local community heated yoga studios or to a waitlist control condition. Participants were recruited between March 2, 2017 and August 5, 2019. Assessments were administered at baseline and biweekly throughout the 8-week primary randomization period. Each study visit included measurement of vital signs, clinician- and self-rated assessments, and assessment of adverse events. Exit evaluations were administered if participants terminated participation early. Study assessments were administered by blinded raters.

Randomization used a permuted-block design sequence with blocks of 2 and 4. The randomization schedule was stored in a locked file, and randomization status was concealed from blinded clinicians. Unblinded study coordinators assigned participants using the specified sequence.

### Participants

Adults with moderate-to-severe depressive symptoms were enrolled via online advertisements through the Massachusetts General Hospital Depression Clinical and Research Program (DCRP). Heated yoga sessions were conducted at two yoga studios in the Boston area.

### Inclusion and exclusion criteria

Inclusion criteria were age 18 to 60 years, current depression indicated by clinician-rated Inventory of Depressive Symptomatology (IDS-CR) score > 23, English language proficiency, and willingness to maintain current medication and exercise regimens during the study. Exclusion criteria included neurological disorders that would impact participation; a history of bipolar disorder, psychotic disorder, eating disorder, or substance use disorder with less than 12 months of remission before screening; medical conditions affecting safety (e.g., unstable/contraindicated cardiac risk factors or heat intolerance); antidepressant medication initiated within 8 weeks or dose change within 4 weeks prior to screening; current individual or group psychotherapy established for under 3 months; and more than 6 hours of yoga or other mind-body practices (e.g., tai chi, meditation) within 6 months prior to screening. A comprehensive list of inclusion and exclusion criteria is provided in the primary trial report [1].

### Intervention

The intervention was Bikram (“heated”) yoga, a standardized 90-minute sequence of 26 postures, bracketed at the beginning and end with breathing exercises, performed in a room heated to 105°F (40.5°C) with approximately 40% relative humidity [36]. Yoga instructors were Original Hot Yoga certified. The instruction method was based on verbal guidance. The intervention was conducted at two community heated yoga studios in the Boston area.

Participants were prescribed a minimum of two classes per week for 8 weeks and could interchangeably use either studio.

Before starting, each participant completed a 50-minute preparatory session with the principal investigator (PI) or designee on strategies to adapt to exercising in a heated environment (e.g., hydration, when to eat a meal, what to wear, how to breathe) and to promote consistent attendance (e.g., explore any barriers to participation, worries or concerns, questions).

To assess fidelity to the yoga protocol, the yoga research liaison, a senior Original Heated Yoga instructor, dropped in on at least 10% of classes offered to participants.

### Waitlist control condition

The waitlist control followed the same assessment schedule as the heated yoga group. Waitlist participants were asked not to modify current treatments during the waitlist period and were offered the same heated yoga intervention after their 8-week control period.

### Measures

#### Clinical information (Psychiatric History Form)

An instrument adapted from the Structured Clinical Interview for DSM-IV Axis I Disorders, Research Version, Non-Patient Edition (SCID-I/NP) was used to collect demographic information, age of onset of depressive illness, psychiatric history, concurrent treatment, and treatment history [37].

#### Depression severity (IDS-CR)

The 30-item clinician-rated Inventory of Depressive Symptomatology (IDS-CR) assessed depressive symptom severity [38]. Scores range from 0 to 84, with higher scores indicating greater severity; a score above 23 is generally considered indicative of moderate-to-severe depressive symptoms.

#### Rumination (RRS)

The 22-item Rumination Response Scale (RRS) assesses responses to depressed mood, including brooding and reflection, on a 4-point Likert scale; higher scores indicate more rumination [39].

#### Mindfulness (FFMQ)

The Five Facet Mindfulness Questionnaire (FFMQ) is a item scale measuring: observing, describing, acting with awareness, nonjudging, and nonreactivity on a 5-point Likert scale; higher scores indicate greater mindfulness [29].

### Statistical analysis

#### Software and overview

Analyses were conducted in R 4.2.2, and causal mediation models were implemented using the `mediation` package [40]. Regression coefficients are reported as unstandardized estimates (*B*) throughout.

#### Participant inclusion criteria

Participants were included if they completed at least one assessment post-baseline, and participants in the yoga condition were additionally required to have attended at least one heated yoga class. Because the `mediation` package requires complete cases, efforts were made to minimize missing Week 4 and Week 8 data: when available, Week-6 values were used to replace missing Week-8 values, and Week-2 values were used to replace missing Week-4 values for mindfulness, rumination, and depressive symptom severity. Participant inclusion criteria for each mediation model were as follows:

1. **Rumination model**: IDS-CR data at baseline and Week 8, and RRS data at baseline and Week 4.
2. **Mindfulness model**: IDS-CR data at baseline and Week 8, and FFMQ data at baseline and Week 4.

All outcome measures (IDS-CR, RRS, FFMQ) were administered at baseline and biweekly throughout the 8-week period (Weeks 2, 4, 6, and 8). For the mediation models, Week 4 values served as the mediator timepoint (with Week 2 values substituted for missing Week 4 observations), and Week 8 values served as the outcome timepoint (with Week 6 values substituted for missing Week 8 observations). The selection of Week 4 as the mediator timepoint and Week 8 as the outcome reflects a temporally ordered framework consistent with prior mediation studies in behavioral interventions, though we acknowledge that more frequent assessments would strengthen causal inference.

Values from the immediately preceding timepoint were carried forward (LOCF) when Week 4 or Week 8 data were unavailable, resulting in 8 substitutions (14.3%) for RRS at Week 4, 6 substitutions (10.0%) for FFMQ at Week 4, and 3 substitutions (4.6%) for IDS-CR at Week 8. These substitutions preserved temporal ordering (mediator preceding outcome) while maximizing sample retention for mediation modeling. LOCF may underestimate variability and bias estimates toward the null; results should therefore be interpreted conservatively given potential attenuation of variance. Patterns of missingness (e.g., MCAR vs. MAR) were not formally tested. Multiple imputation or Bayesian approaches would be preferable and are a priority for future analyses.

The sample size for this secondary analysis was determined by the parent trial (*N* = 80), which was powered to detect between-group differences in IDS-CR scores [1]. A formal a priori power analysis for the mediation models was not conducted.

#### Baseline group comparisons

Group differences in baseline demographic and clinical characteristics were examined using independent-samples *t* tests (for continuous variables) and Fisher’s exact tests (for categorical variables) within each analytic sample.

#### Mediation models

For mediation, we first modeled the mediator at Week 4 (rumination or mindfulness) adjusting for the corresponding baseline mediator scores. We then modeled IDS-CR at Week 8 including the mediator at Week 4, baseline IDS-CR, randomization arm (heated yoga vs. waitlist), and the arm-by-mediator interaction. Average Causal Mediation Effects (ACMEs) by arm, the arm-by-mediator interaction, and total effects were estimated using 10,000 bootstrap simulations with bias-corrected confidence intervals. Sensitivity to residual confounding between mediator and outcome models was assessed with the `medsens` function, which calculates the degree of residual correlation that would shift the ACME confidence interval to include zero. Sensitivity analyses additionally adjusted for baseline covariates that differed significantly by randomization arm.

Regression-based path estimates (Table 2) are reported to provide interpretable effect size information for individual paths, consistent with traditional path-analytic approaches [41]. However, the definitive test of mediation is the ACME bootstrapping framework (Table 3), which has greater statistical power and does not assume a normal sampling distribution for the indirect effect [42].

No correction for multiple comparisons was applied across the two mediator models, as both were prespecified secondary outcomes. The ACME bootstrapping framework estimates arm-specific indirect effects simultaneously; bootstrap resamples were not separately stratified by arm.

These mediation analyses were planned prior to unblinding.

### Sensitivity analyses

Sensitivity analyses examined the degree to which results were resilient to violations of the sequential ignorability assumption, a core assumption of causal mediation analysis requiring that there be no unmeasured confounders of the mediator-outcome relationship conditional on treatment assignment and observed covariates. Specifically, we estimated the percentage of variability that could be shared between residuals of the outcome and mediator models before the observed ACME would no longer differ significantly from zero. Although definitive cut-offs do not exist for these sensitivity analyses, absolute values of *ρ* (rho) exceeding 0.4 are generally considered to indicate that findings are resilient to substantial unmeasured confounding [43].

## Results

The primary depression findings from this RCT were previously published [1] for the 65 participants (heated yoga = 33; waitlist = 32) included in the modified-intent-to-treat analysis (Fig 2). These participants attended an average of 10.3 (± 7.1) total classes over the 8-week intervention period. The mediation analyses were conducted in the subset of participants with complete IDS-CR and mediator data at the required timepoints, as outlined in the Statistical analysis section. Fifty-six participants contributed to the rumination model (heated yoga = 26; waitlist = 30), and sixty participants contributed to the mindfulness model (heated yoga = 28; waitlist = 32). No statistically significant differences in baseline sociodemographic or clinical characteristics were observed between groups (Table 1).

**Table 1.** Participant characteristics at baseline for analytic samples (rumination and mindfulness).

|  | Rumination analyses |  |  | Mindfulness analyses |  |  |
| --- | --- | --- | --- | --- | --- | --- |
|  | HY (*n* = 26) | WL (*n* = 30) | *p* | HY (*n* = 28) | WL (*n* = 32) | *p* |
| **Race, % (*n*)** |  |  |  |  |  |  |
| Asian | 3 (1) | 3 (1) |  | 7 (2) | 7 (2) |  |
| Black | 14 (4) | 13 (4) |  | 14 (4) | 13 (4) |  |
| White | 66 (19) | 68 (20) |  | 69 (20) | 66 (21) |  |
| Multiple | 7 (2) | 10 (3) |  | 7 (2) | 10 (3) |  |
| Unknown | 0 (0) | 3 (1) |  | 0 (0) | 6 (2) |  |
| Hispanic, % (yes) | 8 (2) | 17 (5) | .430 | 7 (2) | 16 (5) | .430 |
| Gender, % (female) | 77 (20) | 93 (28) | .130 | 75 (21) | 94 (30) | .070 |
| Age, y, mean (SD) | 30.27 (8.97) | 33.26 (12.31) | .300 | 31.85 (10.67) | 33.13 (11.93) | .670 |
| **Education, % (*n*)** |  |  |  |  |  |  |
| Less than college degree | 12 (3) | 10 (3) |  | 11 (3) | 6 (3) |  |
| College degree or more | 88 (23) | 90 (27) |  | 89 (25) | 94 (29) |  |
| **Marital status, % (*n*)** |  |  |  |  |  |  |
| Married | 27 (7) | 17 (5) | .520 | 29 (8) | 22 (7) | .570 |
| Not married | 73 (19) | 83 (25) |  | 71 (20) | 78 (25) |  |
| Baseline IDS-CR, mean (SD) | 35.97 (8.66) | 34.47 (6.81) | .480 | 36.37 (9.34) | 34.38 (6.68) | .350 |
| Baseline RRS, mean (SD) | 60.65 (12.97) | 54.10 (9.44) | .050 | 60.65 (12.97) | 54.10 (9.44) | .050 |
| Baseline FFMQ, mean (SD) | 114.54 (19.24) | 115.47 (19.55) | .860 | 114.11 (19.06) | 114.88 (19.05) | .880 |

HY = heated yoga; WL = waitlist; IDS-CR = Inventory of Depressive Symptomatology–Clinician Rated; RRS = Rumination Response Scale; FFMQ = Five Facet Mindfulness Questionnaire; SD = standard deviation. Demographic data were previously reported in Nyer et al. [1]. *p*-values from independent-samples *t*-tests (continuous) and Fisher’s exact tests (categorical); *p*-values reported to 3 decimal places.

**Fig 1.** Conceptual mediation frameworks. Panel A illustrates rumination as the mediator, and Panel B illustrates mindfulness as the mediator. In each model, we test whether the intervention (heated yoga vs. waitlist) reduces depression severity at Week 8 through both direct and indirect pathways. The *direct effect* (*c*^*′*^) is the effect of treatment on depression while controlling for the mediator. The *indirect effect* is the product of the effect of treatment on the mediator (Path *a*) and the effect of the mediator on depression (Path *b*). For rumination, the indirect effect is *a*_1_*b*_1_, and for mindfulness it is *a*_2_*b*_2_. The *total effect* of treatment on depression (*c*) is the sum of the direct and indirect effects (*c* ≈*c*^*′*^ + *ab*). This framework motivates the study hypothesis that changes in rumination and/or mindfulness at Week 4 may mediate the depressive symptom reduction of heated yoga observed at Week 8.

**Fig 2.** CONSORT flow diagram. CONSORT flow diagram for the parent randomized controlled trial of heated yoga (HY) versus waitlist control for adults with moderate-to-severe depression [1]. Of 110 individuals assessed for eligibility, 80 were randomized (40 HY; 40 waitlist). Sixty-five participants (HY *n* = 33; waitlist *n* = 32) were included in the modified intent-to-treat (MITT) analysis of the primary depression endpoint. The present prespecified secondary mediation analyses were conducted in the subsets with complete mediator and outcome data: *n* = 56 for the rumination model (HY *n* = 26; waitlist *n* = 30) and *n* = 60 for the mindfulness model (HY *n* = 28; waitlist *n* = 32). MITT = modified intent-to-treat.

As described in the *Participant inclusion criteria*, to maximize data completeness, when Week-4 or Week-8 data were not available, values from the immediately preceding timepoint (Week 2 or Week 6, respectively) were carried forward, resulting in 8 replacements (14.3%) for RRS at Week 4, 6 replacements (10.0%) for FFMQ at Week 4, and 3 replacements (4.6%) for IDS-CR at Week 8.

Participants randomized to heated yoga showed significantly greater improvements than waitlist controls. Specifically, depressive symptoms decreased from 36.86 to 17.93 in the heated yoga group (Δ = − 18.93, *p* < .001), whereas waitlist participants decreased from 34.38 to 29.63 (Δ = − 4.75, n.s.). Mindfulness increased from 114.09 to 122.66 in the heated yoga group (Δ = +8.57, *p* < .001), while the waitlist group declined from 114.88 to 110.79 (Δ = − 4.09, n.s.). Rumination decreased from 60.52 to 50.97 in the heated yoga group (Δ = − 9.55, *p* < .01), compared with little change in the waitlist group (Δ = +0.52, n.s.) (Fig 3).

**Fig 3.** Changes in depression, mindfulness, and rumination scores over time by group. Mean scores are shown for the heated yoga (HY) and waitlist control groups across two timepoints. Error bars indicate 95% confidence intervals. Panel A (left): depression score from baseline to Week 8 (lower scores indicate improvement). Panel B (middle): mindfulness from baseline to Week 4 (higher scores indicate improvement). Panel C (right): rumination from baseline to Week 4 (lower scores indicate improvement). Asterisks indicate statistically significant within-group change for the heated yoga group relative to baseline (*** *p* < 0.001, ** *p* < 0.01).

### Mediation analyses

Randomization to heated yoga was associated with both a significant decrease in rumination (*B* = − 8.58, *SE* = 2.50, *p* < .01) and a significant increase in mindfulness (*B* = 12.18, *SE* = 2.92, *p* < .001) at study midpoint, after controlling for baseline metrics.

The regression models indicated that the relationships between Week-4 rumination or mindfulness and Week-8 depression severity were similar in the heated yoga and waitlist groups, after controlling for baseline depression, IDS-CR (Table 2). The interaction between mindfulness and treatment condition was not statistically significant (*p >* .10). However, the mediation models (ACME framework) provide the definitive test of indirect effects [43]. Notably, individual path coefficients (Table 2) were not statistically significant, which can occur in mediation analyses where the indirect effect is detectable via bootstrapping despite non-significant component paths.

**Table 2.** Effect estimates of mediator *×* treatment on depressive symptoms at endpoint.

| Model 1: Rumination as mediator |  |  |  |  |  |
| --- | --- | --- | --- | --- | --- |
|  | *B* | SE | *t* | df | *p* |
| Rumination (Week 4) | 0.14 | 0.15 | 0.90 | 51 | .373 |
| Heated yoga treatment | −18.47 | 11.49 | −1.61 | 51 | .114 |
| Rumination × Treatment | 0.14 | 0.22 | 0.65 | 51 | .520 |
| Baseline IDS-CR | 0.68 | 0.18 | 3.69 | 51 | <.001 |
| *F*-statistic: $F(4, 51) = 11.20, p < .001$ | | | | | |
| Model 2: Mindfulness as mediator |  |  |  |  |  |
|  | *B* | SE | *t* | df | *p* |
| Mindfulness (Week 4) | −0.01 | 0.09 | −0.10 | 55 | .919 |
| Heated yoga treatment | 14.18 | 14.41 | 0.98 | 55 | .329 |
| Mindfulness × Treatment | −0.21 | 0.12 | −1.75 | 55 | .086 |
| Baseline IDS-CR | 0.71 | 0.16 | 4.48 | 55 | <.001 |
| *F*-statistic: $F(4, 55) = 15.05, p < .001$ | | | | | |

*B* = unstandardized regression coefficient; SE = standard error; *t* = *t*-statistic; df = degrees of freedom. Week 4 = study midpoint; endpoint = Week 8. IDS-CR = Inventory of Depressive Symptomatology–Clinician Rated; HY = heated yoga; WL = waitlist. Coefficients and SEs reported to 2 decimal places; *p*-values to 3 decimal places.

Assignment to heated yoga was associated with a significant reduction in depressive symptoms in both models, as reflected in the total effect estimates (Model 1: − 13.10, 95% CI [− 18.50, − 7.71]; Model 2: 11.74, 95% CI [− 17.04, − 7.58]; Table 3). In the rumination model (Model 1), the ACME estimates for both the waitlist (− 1.19, 95% CI [− 5.39, 0.97]) and heated yoga groups (− 2.41, 95% CI [− 6.28, 0.43]) were not statistically significant, and the rumination *×* treatment interaction (− 1.22, 95% CI [− 5.18, 3.05]) was also not significant, indicating that reductions in rumination were not statistically consistent with mediating the effect of heated yoga on depressive symptoms. In the mindfulness model (Model 2), the ACME estimate for the heated yoga group had a 95% confidence interval that did not include zero (− 2.71, 95% CI [− 5.42, − 0.99]), and the mindfulness *×* treatment interaction likewise had a confidence interval excluding zero (− 2.59, 95% CI [− 6.12, 0.07]), indicating that increases in mindfulness were statistically consistent with partially mediating the depressive symptom reduction associated with heated yoga.

**Table 3.** Mediation effect estimates of heated yoga treatment on depressive symptoms.

| **Model 1: Rumination as mediator** |  |  |
| --- | --- | --- |
| *Outcome: depressive symptom severity at study endpoint* |  |  |
|  | Estimate | 95% CI |
| *Average Causal Mediation Effect (ACME)* |  |  |
| Waitlist | −1.19 | [−5.39, 0.97] |
| Heated yoga treatment | −2.41 | [−6.28, 0.43] |
| Rumination × HY treatment | −1.22 | [−5.18, 3.05] |
| Total effect | −13.10 | [−18.50, −7.71]* |
| *Sensitivity to pre-treatment confounding ($\rho$)* |  |  |
| Waitlist |  | 0.20 |
| Heated yoga treatment |  | 0.50 |
| **Model 2: Mindfulness as mediator** |  |  |
| *Average Causal Mediation Effect (ACME)* |  |  |
| Waitlist | −0.12 | [−2.22, 2.36] |
| Heated yoga treatment | −2.71 | [−5.42, −0.99]* |
| Mindfulness × HY treatment | −2.59 | [−6.12, −0.07]* |
| Total effect | −11.74 | [−17.04, −7.58]* |
| *Sensitivity to pre-treatment confounding ($\rho$)* |  |  |
| Waitlist |  | 0.00 |
| Heated yoga treatment |  | −0.60 |

ACME = Average Causal Mediation Effect; CI = confidence interval; HY = heated yoga; WL = waitlist; IDS-CR = Inventory of Depressive Symptomatology–Clinician Rated; RRS = Rumination Response Scale; FFMQ = Five Facet Mindfulness Questionnaire. Negative estimates indicate lower depressive symptoms. 95% confidence intervals are bias-corrected bootstrap intervals based on 10,000 simulations. \*95% CI does not include zero. Estimates and $\rho$ values reported to 2 decimal places.

### Sensitivity analyses results

In our models, the ACME for the heated yoga condition was resilient to unmeasured confounding in both Model 1 (*ρ* = 0.5) and Model 2 (*ρ* = − 0.6), indicating that a very strong correlation between unobserved confounders of the mediator-outcome relationship would be required to reduce the mediation effect to zero, regardless of whether the ACME itself reached statistical significance (Table 3).

## Discussion

This study provides evidence consistent with mindfulness as a key behavioral mechanism through which a multicomponent lifestyle intervention (heated yoga) reduces depressive symptoms. In a prespecified secondary analysis of an RCT that demonstrated the efficacy of HY for moderate-to-severe depression [1], causal mediation analysis identified mindfulness as a statistically significant mediator of the treatment effect, while rumination did not emerge as a mediator. These findings advance our understanding of how behavioral lifestyle interventions exert antidepressant effects and have implications for intervention design and personalization.

### Mindfulness as a behavioral mechanism

The finding that increased mindfulness mediates depressive symptom reductions is consistent with a growing body of evidence linking mindfulness-based processes to depression outcomes. Mindfulness-based interventions have repeatedly demonstrated efficacy in reducing depressive symptoms [25, 44, 45], and non-heated yoga interventions that enhance mindfulness have similarly shown antidepressant effects [15, 31, 46, 47]. The present findings extend this literature by demonstrating that mindfulness can be cultivated through a physically demanding, heat-based behavioral intervention, not only through formal meditation or standard yoga.

From a behavioral medicine perspective, mindfulness may function as a regulatory mechanism that alters how individuals engage with negative cognitive and affective experiences. Rather than directly changing thought content, mindfulness may promote decentering, allowing individuals to observe distressing thoughts without habitual reactive engagement. In the context of HY specifically, the practice of maintaining attentional focus on physical sensations during sustained thermal discomfort may serve as a naturalistic form of interoceptive exposure training, building capacity for non-reactive awareness that generalizes to emotional distress outside the practice setting. It is possible that physiological stress induced by heat enhances attentional engagement with interoceptive signals, thereby amplifying mindfulness-related processes.

### Rumination: downstream change rather than independent mechanism

Although rumination scores decreased significantly in the heated yoga group (Fig 3), the association between rumination at midpoint and depressive symptoms at endpoint was not statistically significant after adjusting for baseline depression severity (see Rumination *×* Treatment in Table 2). This pattern suggests that reductions in rumination may reflect downstream changes associated with symptom improvement rather than an independent mediating pathway. This interpretation is consistent with prior findings from Nolen-Hoeksema [32] and Broderick and Korteland [48], who reported that rumination is closely tied to depression severity rather than functioning as an independent mechanism of change.

The contrast between mindfulness and rumination results is noteworthy from a mechanistic standpoint: it suggests that HY may primarily operate by building new regulatory capacities (present-moment awareness, non-reactivity) rather than by directly disrupting perseverative cognitive processes. This distinction has practical implications for intervention design, suggesting that behavioral treatments emphasizing attentional training and interoceptive awareness may be more effective than those targeting rumination reduction directly.

### Regression path vs. bootstrap mediation discrepancy

The interaction between mindfulness and treatment condition was not statistically significant in the regression model testing the association between midpoint mindfulness and endpoint depression (Table 2). This may reflect limited power to detect cross-level interactions in a modest sample. Mediation analyses using the ACME framework provide a more appropriate test of indirect effects, directly estimating the extent to which changes in mindfulness account for reductions in depressive symptoms (Table 3). The non-significant interaction should be interpreted in the context of the mediation findings, which provide a more appropriate test of the indirect effect.

### Integration of physiological and behavioral pathways

Heated yoga is a multicomponent behavioral intervention that engages both psychological and physiological processes. Exposure to heat at sufficient intensity and duration can induce whole-body hyperthermia (WBH), a treatment with independent evidence supporting depressive symptom reduction [18, 19]. Depression has been linked with persistent elevations in core body temperature and blunted thermoregulatory cycling [19, 20, 22, 49], and these temperature abnormalities appear to normalize following WBH interventions [19, 50].

Beyond the psychological mechanisms examined here, growing evidence implicates neurobiological and immunological pathways in the antidepressant effects of heat-based interventions. Flux et al. demonstrated that the antidepressant response to whole-body hyperthermia in MDD is associated with acute changes in plasma cytokines, particularly interleukin-6 [51]. A recent meta-analysis further confirmed that yoga-based interventions significantly reduce both anxiety and depressive symptoms [26]. These converging lines of evidence suggest that HY may reduce depression through simultaneous behavioral-psychological and physiological mechanisms, a hypothesis warranting direct investigation with concurrent measurement of both pathways.

Heated yoga superimposes the exogenous physiological stressor of ambient heat upon the endogenous demands of postures, movement, and breath regulation. Unlike the voluntary modulation of physical or respiratory practices, the thermal environment cannot be readily attenuated or avoided during class participation. In this sense, the practice invites participants to remain present with sustained visceral discomfort, a process that parallels behavioral approaches to distress tolerance and exposure-based treatments for emotional disorders.

### Measurement considerations

The FFMQ comprises five facets: observing, describing, acting with awareness, non-judging of inner experience, and non-reactivity to inner experience. Prior psychometric work has noted that the observing subscale shows weaker convergent validity with the other facets in non-meditating samples [29, 52], and may behave differently following yoga versus formal mindfulness practice. Facet-level analyses were not conducted in the present study. Given that facets such as non-reactivity and acting with awareness are theoretically central to tolerating discomfort in heated environments, future work should prioritize facet-level mediation models to identify which specific facets are most responsive to HY.

A further limitation is that the FFMQ was developed for mindfulness meditation populations. Its use in a yoga context, while supported by prior research [29–31], may not fully capture the interoceptive and somatic awareness dimensions cultivated through yoga practice. Future studies should incorporate interoceptive awareness measures (e.g., the Multidimensional Assessment of Interoceptive Awareness) alongside the FFMQ to better characterize the attentional and regulatory processes engaged by HY.

### Strengths

This study has several notable strengths. The randomized, controlled trial design enhances internal validity. The use of blinded outcome assessors administering gold-standard clinician-rated measures reduces potential bias. The sample was also more racially and ethnically diverse than is typically observed in the yoga literature, with 30.8% of participants identifying as non-white [10]. Finally, the intervention was delivered in a community-based setting, increasing the ecological validity of the findings and supporting the generalizability of results to real-world practice.

### Limitations

It is important to note that mediation analysis, even with bootstrapped confidence intervals and sensitivity analyses, cannot establish causality in the absence of experimental manipulation of the mediator. The present findings are best interpreted as evidence consistent with a mediating role of mindfulness, rather than proof of a causal mechanism.

The use of a waitlist control group, while ethically appropriate, is a methodological limitation: waitlist status may carry psychological costs, such as perceived inequity or demoralization, that could inflate apparent between-group differences. Future trials should include an active control condition.

The absence of an active comparator group limits the ability to interpret specific treatment effects and precludes differentiation between the unique benefits of heated yoga and nonspecific therapeutic factors such as expectancy effects and the social interaction from the assessment visits. While there was more racial and ethnic diversity, the sample consisted primarily of female (82%) and college-educated (86%) participants, which may restrict generalizability. The physical and thermal demands of the intervention exclude individuals with certain medical conditions. The study did not assess treatment resistance or include individuals with mild depression or active comorbid psychiatric conditions, limiting conclusions about its utility across the broader spectrum of depressive illness.

Although the temporal ordering is consistent with a mediation framework, the spacing of assessments limits our ability to definitively establish that changes in mindfulness preceded changes in depressive symptoms at a finer timescale. Denser longitudinal assessment would strengthen causal inference in future work.

Causal mediation analysis assumes no unmeasured confounders between the mediator and the outcome (the sequential ignorability assumption). This assumption cannot be empirically verified; unmeasured confounders such as expectancy, treatment engagement, or concurrent life events could bias the ACME estimates.

Both mediators were assessed via self-report, which is susceptible to response bias and recall error. Future research should complement self-report with ecological momentary assessment or physiological measures (e.g., heart rate variability as a mindfulness index, perseverative cognition indices for rumination).

The study may have been underpowered to detect a rumination mediation effect if one exists; this possibility cannot be excluded.

### Future directions

To establish the clinical utility of heated yoga, future research should prioritize adequately powered multi-site trials. Direct comparisons between heated and non-heated yoga are needed to determine whether heat confers additional benefits for depressive symptoms, particularly given promising findings for whole body hyperthermia as a standalone treatment for depression [18, 20]. Evaluating heated yoga against active comparators, such as established psychotherapies or pharmacological treatments, would help clarify whether its effects are specific or due to an expectancy effect. From a clinical and public health perspective, behavioral lifestyle interventions such as yoga may represent scalable, community-deliverable, non-pharmacologic strategies for targeting core depressive processes. Unlike psychotherapy, which requires trained therapists and often involves lengthy wait times, community-based yoga classes are widely available and may serve as accessible adjunctive treatments. These findings suggest that interventions which cultivate mindfulness, whether through structured MBIs or embodied behavioral practices, may represent a promising class of behavioral treatments for depression.

## Conclusion

In this RCT, a multicomponent behavioral lifestyle intervention (heated yoga) reduced depressive symptoms, and this effect was statistically consistent with mediation by increases in mindfulness but not by reductions in rumination. These findings identify mindfulness as a key behavioral mechanism through which lifestyle interventions may exert antidepressant effects, with implications for optimizing behavioral treatments for depression. Heated yoga was associated with clinically meaningful reductions in depressive symptoms even though participants averaged fewer than the prescribed number of sessions, suggesting that the intervention may be effective at doses achievable in real-world community settings. Future research should build on this work through larger-scale studies with active comparators, denser longitudinal assessment, and concurrent measurement of psychological and physiological mechanisms.

## Data Availability

Data cannot be made publicly available due to ethical and legal restrictions imposed by the Mass General Brigham IRB and the terms of participant informed consent for this randomized controlled trial. The dataset contains sensitive clinical information (psychiatric diagnoses, symptom trajectories, demographics) that poses re-identification risk given the modest sample (N = 80). A de-identified minimal dataset sufficient to reproduce the reported findings is available to qualified researchers upon reasonable request, subject to a data-sharing agreement consistent with the IRB-approved protocol. Requests should be directed to the Mass General Brigham Human Research Office, Somerville, MA, USA ( phone: +1-857-282-1900), referencing IRB Protocol 2016P000704 and ClinicalTrials.gov NCT02607514. Analysis code is available from the corresponding author upon request.

## Supporting information

**S1 Fig. Conceptual mediation frameworks**. Diagram of mediation models with rumination (Panel A) and mindfulness (Panel B) as candidate mediators of the heated yoga → depression effect.

**S2 Fig. CONSORT flow diagram**. Participant flow through screening, randomization, allocation, follow-up, and analysis for the parent trial and the present mediation subsamples.

**S3 Fig. Changes in depression, mindfulness, and rumination scores over time by group**. Group means with 95% confidence intervals for IDS-CR, FFMQ, and RRS at the relevant assessment timepoints.

**S1 Appendix. Supplementary materials**. CLARIFY guidelines checklist, CONSORT 2010 checklist, descriptive statistics at all timepoints, and the standardized Bikram 26-posture series.

## Acknowledgments

The authors express gratitude to Kendra Blackett, MPH; Lucas Lambert, HSD; Shelley Cates, MFA; and Pablo Picker, BA, who provided the heated yoga classes free of charge, collected and reported attendance data, and coordinated teacher training through their respective studios (Bikram Yoga Boston, Bikram Yoga Works Boston, Bikram Yoga Cambridge, and Breathe Cambridge). These individuals taught classes for the regularly held community classes and checked participants in to the classes at the front desk; none had contact with the data collected in this study, and none have relevant financial relationships to report. Funding, data availability, and author contribution information is entered directly into the dedicated fields in the PLOS Editorial Manager submission system and will be incorporated into the appropriate sections of the published article during production.

## References

1. Nyer MB, Mason AE, Jain FA, Pedrelli P, Mischoulon D, Fava M. A randomized controlled trial of community-delivered heated yoga versus waitlist control for moderate-to-severe depression: Feasibility, acceptability, and preliminary efficacy. J Clin Psychiatry. 2023;84(6):23m14267. Trial registration: ClinicalTrials.gov NCT02607514. doi:10.4088/JCP.23m14267.

2. World Health Organization. Mental disorders: Key facts; 2022. Accessed September 10, 2025. Available from: https://www.who.int/news-room/fact-sheets/detail/mental-disorders.

3. Cuijpers P, Karyotaki E, Weitz E, Andersson G, Hollon S, van Straten A. The effects of psychotherapies for major depression in adults on remission, recovery and improvement: a meta-analysis. J Affect Disord. 2014;159:118–26.

4. Levkovitz Y, Tedeschini E, Papakostas G. Efficacy of antidepressants for dysthymia: a meta-analysis of placebo-controlled randomized trials. J Clin Psychiatry. 2011;72(4):596–604.

5. Cassano P, Fava M. Tolerability issues during long-term treatment with antidepressants. Ann Clin Psychiatry. 2004;16(1):15–25.

6. Sarris J, O’Neil A, Coulson CE, Schweitzer I, Berk M. Lifestyle medicine for depression. BMC Psychiatry. 2014;14:107. doi:10.1186/1471-244X-14-107.

7. Firth J, Solmi M, Wootton RE, Vancampfort D, Schuch FB, Hoare E, et al. A meta-review of “lifestyle psychiatry”: the role of exercise, smoking, diet and sleep in the prevention and treatment of mental disorders. World Psychiatry. 2020;19(3):360–80. doi:10.1002/wps.20773.

8. Kandola A, Ashdown-Franks G, Hendrikse J, Sabiston CM, Stubbs B. Physical activity and depression: towards understanding the antidepressant mechanisms of physical activity. Neurosci Biobehav Rev. 2019;107:525–39. doi:10.1016/j.neubiorev.2019.09.040.

9. Paolucci EM, Loukov D, Bowdish DME, Heisz JJ. Exercise reduces depression and inflammation but intensity matters. Biol Psychol. 2018;133:79–84. doi:10.1016/j.biopsycho.2018.01.015.

10. Cramer H, Lauche R, Langhorst J, Dobos G. Yoga for depression: a systematic review and meta-analysis. Depress Anxiety. 2013;30(11):1068–83. doi:10.1002/da.22166.

11. Uebelacker L, Broughton M. Yoga for Depression and Anxiety: A Review of Published Research and Implications for Healthcare Providers. Rhode Island Med J. 2016;99(3):20–2.

12. Wu Y, Yan D, Yang J. Effectiveness of yoga for major depressive disorder: A systematic review and meta-analysis. Frontiers in Psychiatry. 2023 Mar;14:1138205. doi:10.3389/fpsyt.2023.1138205.

13. Brinsley J, Schuch F, Lederman O, Girard D, Smout M, Immink MA, et al. Effects of yoga on depressive symptoms in people with mental disorders: a systematic review and meta-analysis. British Journal of Sports Medicine. 2021;55(17):992–1000. Available from: https://bjsm.bmj.com/content/55/17/992. doi:10.1136/bjsports-2019-101242.

14. Prathikanti S, Rivera R, Cochran A, Tungol JG, Fayazmanesh N, Weinmann E. Treating major depression with yoga: A prospective, randomized, controlled pilot trial. PLOS ONE. 2017;12(3):e0173869. Available from: https://doi.org/10.1371/journal.pone.0173869. doi:10.1371/journal.pone.0173869.

15. Schuver K, Lewis B. Mindfulness-based yoga intervention for women with depression. Complement Ther Med. 2016;26:85–91.

16. Uebelacker L, Epstein-Lubow G, Gaudiano B, Tremont G, Battle C, Miller I. Hatha yoga for depression: critical review of the evidence for efficacy, plausible mechanisms of action, and directions for future research. J Psychiatr Pract. 2010;16(1):22–33.

17. Baird SO, Hopkins LB, Medina JL, Rosenfield D, Powers MB, Smits JA. Distress Tolerance as a Predictor of Adherence to a Yoga Intervention: Moderating Roles of BMI and Body Image. Behavior Modification. 2016;40(1-2):199–217. Epub 2015 Nov 2. doi:10.1177/0145445515612401.

18. Janssen C, Lowry C, Mehl M, Allen J, Kelly K, Gartner D, et al. Whole-Body Hyperthermia for the Treatment of Major Depressive Disorder: A Randomized Clinical Trial. JAMA Psychiatry. 2016;73(8):789–95.

19. Mac Giollabhúi N, Lowry CA, Nyer M, Foster SL, Liu RT, Smith DG, et al. The Antidepressant Effect of Whole-Body Hyperthermia Is Associated with the Classical Interleukin-6 Signaling Pathway. Brain, Behavior, and Immunity. 2024 Jul;119:801–6. Epub 2024 Apr 26; Erratum in: Brain Behav Immun. 2025 Mar;125:471. doi: 10.1016/j.bbi.2025.01.005. Available from: https://www.ncbi.nlm.nih.gov/pmc/articles/PMC11670333/. doi: 10.1016/j.bbi.2024.04.040.

20. Hanusch K, Janssen C, Billheimer D, Jenkins I, Spurgeon E, Lowry C, et al. Whole-body hyperthermia for the treatment of major depression: associations with thermoregulatory cooling. Am J Psychiatry. 2013;170(7):802–4.

21. Hanusch KU, Janssen CW. The impact of whole-body hyperthermia interventions on mood and depression – are we ready for recommendations for clinical application? International Journal of Hyperthermia. 2019;36(1):573–81. Available from: https://doi.org/10.1080/02656736.2019.1612103. doi:10.1080/02656736.2019.1612103.

22. Mason AE, Chowdhary A, Hartogensis W, Siwik CJ, Lupesko-Persky O, Pandya LS, et al. Feasibility and acceptability of an integrated mind-body intervention for depression: whole-body hyperthermia (WBH) and cognitive behavioral therapy (CBT). International Journal of Hyperthermia. 2024;41(1):2351459. Available from: https://doi.org/10.1080/02656736.2024.2351459. doi:10.1080/02656736.2024.2351459.

23. Kabat-Zinn J. Full Catastrophe Living: Using the Wisdom of Your Body and Mind to Face Stress, Pain, and Illness. New York: Delacorte Press; 1990.

24. Kuyken W, Warren FC, Taylor RS, Whalley B, Crane C, Bondolfi G, et al. Efficacy of Mindfulness-Based Cognitive Therapy in Prevention of Depressive Relapse: An Individual Patient Data Meta-analysis From Randomized Trials. JAMA Psychiatry. 2016;73(6):565–74. doi:10.1001/jamapsychiatry.2016.0076.

25. Goldberg SB, Tucker RP, Greene PA, Davidson RJ, Kearney DJ, Simpson TL. Mindfulness-based interventions for psychiatric disorders: A systematic review and meta-analysis. Clin Psychol Rev. 2018;59:52–60. doi:10.1016/j.cpr.2017.10.011.

26. Martínez-Calderon J, Casuso-Holgado MJ, Muñoz-Fernandez MJ, Garcia-Muñoz C, Heredia-Rizo AM. Yoga-based interventions may reduce anxiety symptoms in anxiety disorders and depression symptoms in depressive disorders: a systematic review with meta-analysis and meta-regression. Br J Sports Med. 2023;57(22):1442–9. doi:10.1136/bjsports-2022-106497.

27. Shapiro SL, Carlson LE, Astin JA, Freedman B. Mechanisms of mindfulness. J Clin Psychol. 2006;62(3):373–86. doi:10.1002/jclp.20237.

28. Hölzel BK, Lazar SW, Gard T, Schuman-Olivier Z, Vago DR, Ott U. How Does Mindfulness Meditation Work? Proposing Mechanisms of Action From a Conceptual and Neural Perspective. Perspect Psychol Sci. 2011;6(6):537–59. doi:10.1177/1745691611419671.

29. Baer R, Smith G, Hopkins J, Krietemeyer J, Toney L. Using self-report assessment methods to explore facets of mindfulness. Assessment. 2006;13(1):27–45.

30. Carmody J, Baer RA. Relationships between mindfulness practice and levels of mindfulness, medical and psychological symptoms and well-being in a mindfulness-based stress reduction program. J Behav Med. 2008;31(1):23–33. doi:10.1007/s10865-007-9130-7.

31. Vollbehr NK, Hoenders HJR, Bartels-Velthuis AA, Nauta MH, Castelein S, Schroevers MJ, et al. Mindful Yoga Intervention as Add-on to Treatment as Usual for Young Women With Major Depressive Disorder: Results From a Randomized Controlled Trial. Journal of Consulting and Clinical Psychology. 2022;90(12):925–41. doi:10.1037/ccp0000777.

32. Nolen-Hoeksema S. The role of rumination in depressive disorders and mixed anxiety/depressive symptoms. J Abnorm Psychol. 2000;109(3):504–11. doi:10.1037/0021-843X.109.3.504.

33. Robinson MS, Alloy LB. Negative cognitive styles and stress-reactive rumination interact to predict depression: A prospective study. Cognitive Ther Res. 2003;27(3):275–91. doi:10.1023/A:1023914416469.

34. Moonaz S, Nault D, Cramer H, et al. CLARIFY 2021: explanation and elaboration of the Delphi-based guidelines for the reporting of yoga research. BMJ Open. 2021;11(8):e045812. Available from: https://bmjopen.bmj.com/content/11/8/e045812. doi:10.1136/bmjopen-2020-045812.

35. Schulz KF, Altman DG, Moher D, Group C. CONSORT 2010 Statement: updated guidelines for reporting parallel group randomised trials. BMC Medicine. 2010;8(1):18. Available from: https://bmcmedicine.biomedcentral.com/articles/10.1186/1741-7015-8-18. doi:10.1186/1741-7015-8-18.

36. Nyer M, Hopkins L, Farabaugh A, Nauphal M, Parkin S, McKee M, et al. Community-Delivered Heated Hatha Yoga as a Treatment for Depressive Symptoms: An Uncontrolled Pilot Study. J Altern Complement Med. 2019;25(8):814–23.

37. First M, Gibbon M, Spitzer R, Williams J, Benjamin L. Structured Clinical Interview for DSM-IV Axis I Disorders, Research Version, Non-Patient Edition (SCID-I/NP). New York State Psychiatric Institute; 1997.

38. Rush A, Gullion C, Basco M, Jarrett R, Trivedi M. The Inventory of Depressive Symptomatology (IDS): psychometric properties. Psychol Med. 1996;26(3):477–86.

39. Nolen-Hoeksema S, Morrow J. A prospective study of depression and posttraumatic stress symptoms after a natural disaster: the 1989 Loma Prieta Earthquake. J Pers Soc Psychol. 1991;61(1):115.

40. Tingley D, Yamamoto T, Hirose K, Keele L, Imai K. mediation: R Package for Causal Mediation Analysis; 2014. R package. Available from: https://cran.r-project.org/package=mediation.

41. Baron RM, Kenny DA. The moderator–mediator variable distinction in social psychological research: Conceptual, strategic, and statistical considerations. J Pers Soc Psychol. 1986;51(6):1173–82. doi:10.1037/0022-3514.51.6.1173.

42. Fritz MS, Cox MG, MacKinnon DP. Increasing statistical power in mediation models without increasing sample size. Evaluation and the Health Professions. 2015;38(3):343–66. doi:10.1177/0163278713514250.

43. Imai K, Keele L, Tingley D, Yamamoto T. General approach to causal mediation analysis. Psychological Methods. 2010;15(4):309–34. doi:10.1037/a0020761.

44. Meadows GN, Shawyer F, Enticott JC, Graham AL, Judd F, Martin PR, et al. Mindfulness-based cognitive therapy for recurrent depression: A translational research study with 2-year follow-up. Aust N Z J Psychiatry. 2014 Aug;48(8):743–55. Epub 2014 Mar 4. doi:10.1177/0004867414525841.

45. Saeed SA, Cunningham K, Bloch RM. Depression and anxiety disorders: Benefits of exercise, yoga, and meditation. Am Fam Physician. 2019;99(10):620–7.

46. Miao C, Gao Y, Li X, et al. The effectiveness of mindfulness yoga on patients with major depressive disorder: a systematic review and meta-analysis of randomized controlled trials. BMC Complementary Medicine and Therapies. 2023;23:313. Available from: https://doi.org/10.1186/s12906-023-04141-2. doi:10.1186/s12906-023-04141-2.

47. Medina J, Hopkins L, Powers M, Baird SO, Smits J. The Effects of a Hatha Yoga Intervention on Facets of Distress Tolerance. Cognitive Behaviour Therapy. 2015;44(4):288–300. Epub 2015 May 8. doi:10.1080/16506073.2015.1028433.

48. Broderick PC, Korteland C. A Prospective Study of Rumination and Depression in Early Adolescence. Clinical Child Psychology and Psychiatry. 2004;9(3):383-94. First published online Jul 2004. doi:10.1177/1359104504043920.

49. Mason AE, Kasl P, Soltani S, et al. Elevated Body Temperature Is Associated with Depressive Symptoms: Results from the TemPredict Study. Scientific Reports. 2024;14:1884. doi:10.1038/s41598-024-51567-w.

50. Lowry CA, Flux M, Raison CL. Whole-Body Heating: An Emerging Therapeutic Approach to Treatment of Major Depressive Disorder. Focus (American Psychiatric Publishing). 2018;16(3):259–65. Epub 2018 Jul 16. doi:10.1176/appi.focus.20180009.

51. Flux MC, Smith DG, Allen JJB, Mehl MR, Medrano A, Begay TK, et al. Association of plasma cytokines and antidepressant response following mild-intensity whole-body hyperthermia in major depressive disorder. Transl Psychiatry. 2023;13(1):132. doi:10.1038/s41398-023-02402-9.

52. Williams MJ, Dalgleish T, Karl A, Kuyken W. Examining the factor structures of the Five Facet Mindfulness Questionnaire and the Self-Compassion Scale. Psychological Assessment. 2014;26(2):407–18. doi:10.1037/a0035566.

